# Effect of SARS-CoV-2 vaccination on symptoms from post-acute COVID syndrome: results from the national VAXILONG survey

**DOI:** 10.1101/2021.10.11.21264849

**Authors:** Marc Scherlinger, Luc Pijnenburg, Emmanuel Chatelus, Laurent Arnaud, Jacques-Eric Gottenberg, Jean Sibilia, Renaud Felten

## Abstract

**Introduction:** Few data are available concerning the effect of SARS-CoV-2 vaccination on the persistent symptoms associated with COVID-19, also called long-COVID or post-acute COVID-19 syndrome (PACS).

**Patients and methods:** We conducted a nationwide online survey among adult patients with PACS as defined by symptoms persisting over 4 weeks following a confirmed or probable COVID-19, without any identified alternative diagnosis. Information concerning PACS symptoms, vaccine type and scheme and its effect on PACS symptoms were studied.

**Results:** Six hundred and twenty surveys were completed and 567 satisfied the inclusion criteria and were analyzed. Respondents were 83.4% of women of median age 44 (IQR 25-75: 37-50). Initial infection was proven in 365 patients (64%) and 5.1% had been hospitalized to receive oxygen. 396 patients had received at least one injection of SARS-CoV-2 vaccine at the time of the survey, after a median of 357 [198-431] days following the initially-reported SARS-CoV-2 infection. Among the 380 patients who reported persistent symptoms at the time of SARS-CoV-2 vaccination, 201 (52.8%) reported variation of symptoms following the injection, without difference based on the type of vaccine used. After a complete vaccination scheme, 93.3% (28/30) of initially seronegative patients reported a positive anti-SARS-CoV-2 IgG.

170 PACS patients had not been vaccinated. The most common reasons for postponing SARS-CoV-2 vaccine were a fear of worsening PACS symptoms (55.9%) and the idea that vaccination was contraindicated because of PACS (15.6%).

**Conclusion:** Our study suggests that SARS-CoV-2 vaccination is well tolerated in the majority of PACS patients and has good immunogenicity. Disseminating these reassuring data might prove crucial to increase vaccine coverage in patients with PACS.

## Introduction

It is estimated that 10% of patients infected with SARS-CoV-2 will continue to experience debilitating symptoms at least 12 weeks after their initial infection [1], a condition called long-COVID or post-acute COVID syndrome (PACS). The etiopathogenesis of PACS is uncertain and probably multifactorial, including viral- or immune-mediated organ injury, neurological involvement, dysautonomia, physical deconditioning and psychological burden [2,3]. These uncertainties have caused PACS patients to fear adverse effects from vaccination, prompting some not to take part in the current campaign [4]. On the other hand, viral persistence due to defective anti-viral immunity has been hypothesized to account for PACS [5], paving the way to use SARS-CoV-2 vaccination as a treatment to restore viral immunity and improve symptoms burden. Indeed, preliminary data suggest that SARS-CoV-2 vaccination could improve PACS symptoms [6]. The aim of this study was to evaluate the impact of SARS-CoV-2 vaccination on PACS burden.

### Patients and methods

We conducted an online survey (Google Form^®^) among French-speaking adults recruited through social media platform (*i*.*e*., Twitter^®^, Facebook^®^) and patient association (*i*.*e*., Après-J20).

The survey was anonymous, approved by an independent ethics committee (CE-2021-106) and all patients provided informed consent.

Inclusion criteria were the definition of PACS by the French Haute Autorité de Santé [7]: a reported viral illness with a probable or confirmed COVID-19 diagnosis, persistent symptoms lasting > 4 weeks and the lack of an alternative diagnosis to explain the presentation.

The severity of a wide set of symptoms before and after vaccination was evaluated using a previously validated symptom set [8]. Information about the type of vaccine used or the reason for non-vaccination were evaluated.

Quantitative data are reported as median with interquartile range [IQR 25-75] and qualitative results as percentage. Quantitative data were compared using Student’s t-test, and qualitative data using Chi2 test. Statistical analysis was conducted using JMP Software 14.0 (SAS institute, Cary, USA). A p-value < 0.05 was considered statistically significant.

## Results

Six hundred and twenty patients completed the survey between August 3^rd^ and 17^th^ 2021, and 567 (91.5%) satisfied the inclusion criteria and were included in the analysis (**Figure 1**). Respondents were 83.4% women with a median age of 44 years (IQR 25-75: 37-50). Initial infection was proven (with either RT-PCR, CT-scanner, serology or antigen test) in 365 patients (64%) and 5.1% had been hospitalized to receive oxygen. 396 patients had received at least one injection of SARS-CoV-2 vaccine at the time of the survey, after a median of 357 [198-431] days following the initially-reported SARS-CoV-2 infection. Among them, 255 (64.4%) had a complete vaccination scheme, including 142 with 2 doses and 113 with 1 dose and prior positive RT-PCR or serology. Other patient characteristics are shown in **Supplementary Table 1**.

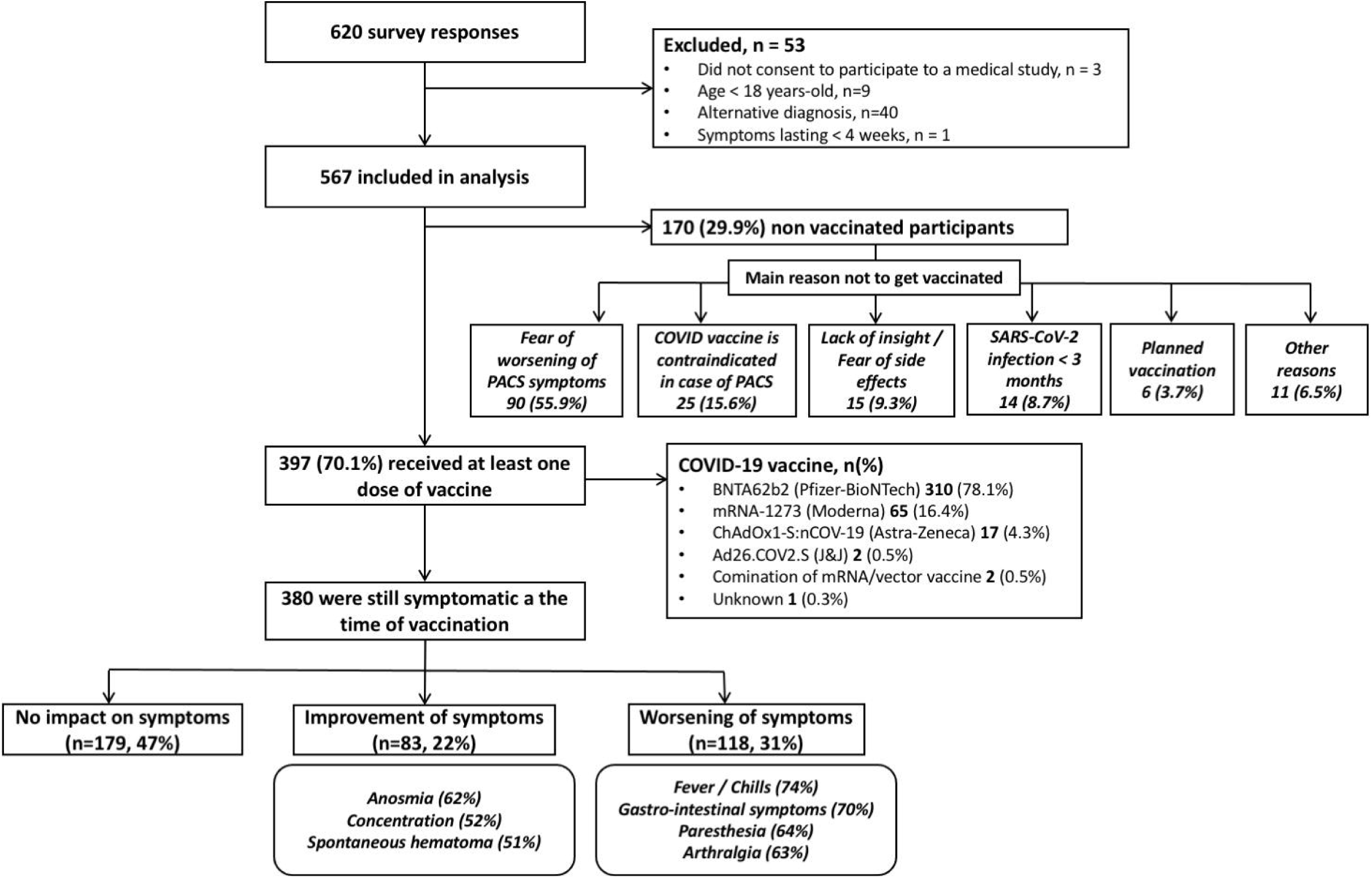

Among the 380 patients who reported persistent symptoms at the time of SARS-CoV-2 vaccination, 201 (52.8%) reported variation of symptoms following the injection. The impact of SARS-CoV-2 vaccination on PACS was not different depending the vaccine used (p=0.60). A worsening of symptom severity was reported by 117 patients (31% of vaccinated PACS patients) and were mostly represented by fever/chills (74%), gastro-intestinal symptoms (70%), paresthesia (64%) and arthralgia (63%). Conversely, symptom improvement was reported by 83 (21.8%) and included anosmia (62%) and brain fog (51%). Vaccine impact on each symptom is shown in **supplementary Figure 1**. Vaccine impact on PACS symptoms lasted more than 2 weeks in 67.8% of patients. The frequency of global improvement following SARS-CoV-2 vaccination was similar between virologically-confirmed and non-virologically confirmed PACS patients: 20.2% (48/238) *vs*. 24.3% (34/140), respectively, p=0.35). However, non-virologically-confirmed PACS patients were more likely to report symptom worsening following vaccination compared to the others 41.4% (58/140) *vs*. 24.8% (59/238) (p <0.001). After a complete vaccination scheme, 93.3% (28/30) of initially seronegative patients reported a positive anti-SARS-CoV-2 IgG.

At the time of the study, 170 PACS patients had not been vaccinated. The characteristics of these patients were similar to vaccinated ones, except for a shorter delay between COVID-19 and survey completion (**supplementary Table 1**). The most common reasons for postponing SARS-CoV-2 vaccine were a fear of worsening PACS symptoms (55.9%) and the idea that vaccination was contraindicated because of PACS (15.6%).

## Discussion

Our study suggests that more than two third of patients with PACS may be vaccinated against SARS-CoV-2 without worsening of their symptoms. Fever/chills and gastro-intestinal symptoms, were the most frequently reported worsened symptoms, but are also commonly reported after SARS-Cov-2 vaccination in the general population [9]. Conversely, one out of 5 patients reported an improvement of their symptoms, mainly brain fog and anosmia which have been associated with disability in PACS. As previously shown [3], more than 90% of PACS patients report fluctuation of their symptoms which may account for some of the findings. Interestingly, the vast majority of patients (93.3%) who had a post-vaccinal serology developed detectable anti SARS-CoV-2 IgG, suggesting normal immunogenicity of the vaccine in this PACS population. The characteristics of the vaccinated and non-vaccinated PACS populations were similar, except for the delay since the initial infection which was shorter in the non-vaccinated population (**supplementary table 1**). This was expected since French health authorities have recommended to postpone SARS-CoV-2 vaccination at least 3 months after COVID-19. The willingness to get vaccinated against SARS-CoV-2 in patients with PACS was limited by the fear of side-effects, the belief that it is contraindicated in PACS and the paucity of available data. Our results suggest that not only SARS-CoV-2 vaccination is well tolerated in the majority of PACS patients, but also that its immunogenicity appears to be normal in this population.

Our study has limitations. First, the recruitment was conducted using social media platforms which could select a younger population. However, the age distribution of the PACS population was similar to that previously reported [3,10]. Second, the vast majority of patients received a mRNA-based vaccine, limiting the generalization of our findings to vector- or antigen-based vaccines. However, these data reflect the vaccines generally used in Europe or in the United States in this young population.

In conclusion, our study suggests that SARS-CoV-2 vaccination is well tolerated in the majority of PACS patients and has good immunogenicity. Disseminating these reassuring data might prove crucial to increase vaccine coverage in patients with PACS.

## Data Availability

All data produced in the present study are available upon reasonable request to the authors.

