## Supplementary Table 1 for "Effect of SARS-CoV-2 vaccination on symptoms from post-acute COVID syndrome: results from the national VAXILONG survey"

|  | Total PACS population  (n = 567) | Vaccinated population  (n = 397) | Non-vaccinated population  (n = 170) | p-value  vaccinated vs.  Non-vaccinated |
| --- | --- | --- | --- | --- |
| Female sex, % (n) | 83.4% (473) | 85.9% (327) | 82.4% (146) | *ns* |
| Age, median (IQR) | 44 (37-50) | 44 (37-50) | 42 (36-49) | *ns* |
| COVID-19 severity   - Home-care - Hospitalized with oxygen therapy   - Intensive care unit | 94.9% (538)  5.1 (25)  0.7% (4) | 95.3% (376)  4.7 (18)  0.8% (3) | 94.7% (162)  5 3 (7)  0.6% (1) | *ns* |
| Confirmed COVID-19, % (n)   - Positive RT-PCR - Positive lung CT - Positive serology - Positive antigen test | 64.4% (365)  45% (255)  22.8% (129)  32.3% (183)  8.6% (49) | 63% (250)  43.1% (171)  22.2% (88)  30.2% (120)  8.1% (32) | 67.7% (115)  49.4% (84)  24.1% (41)  37.1% (63)  10% (17) | *ns* |
| Time since initial COVID-19, days, median (IQR) | 475 (261-506) | 483 (266-506) | 325 (180-507) | **0.0066** |
| Number of persisting symptoms, median (IQR) | 12 (9-15) | 12 (9-15) | 13 (10-15) | *ns* |
| Professional activity   - Unchanged - Adapted to PACS - Interrupted | 45% (255)  18% (102)  37% (210) | 47.4% (188)  19.4% (77)  33.3% (132) | 39.4% (67)  14.7% (25)  45.9% (78) | **0.016** |
| Vaccination   - Time since initial infection - Doses   - One   - Two | - | 357 [198-431]  64.2% (255)  35.8% (142) | -  - | N/A |

**Supplementary Table 1: Characteristics from the included population.**

Lung CT, lung computed tomography; PACS, post-acute COVID-19 syndrome; RT-PCR, reverse transcriptase polymerase chain reaction

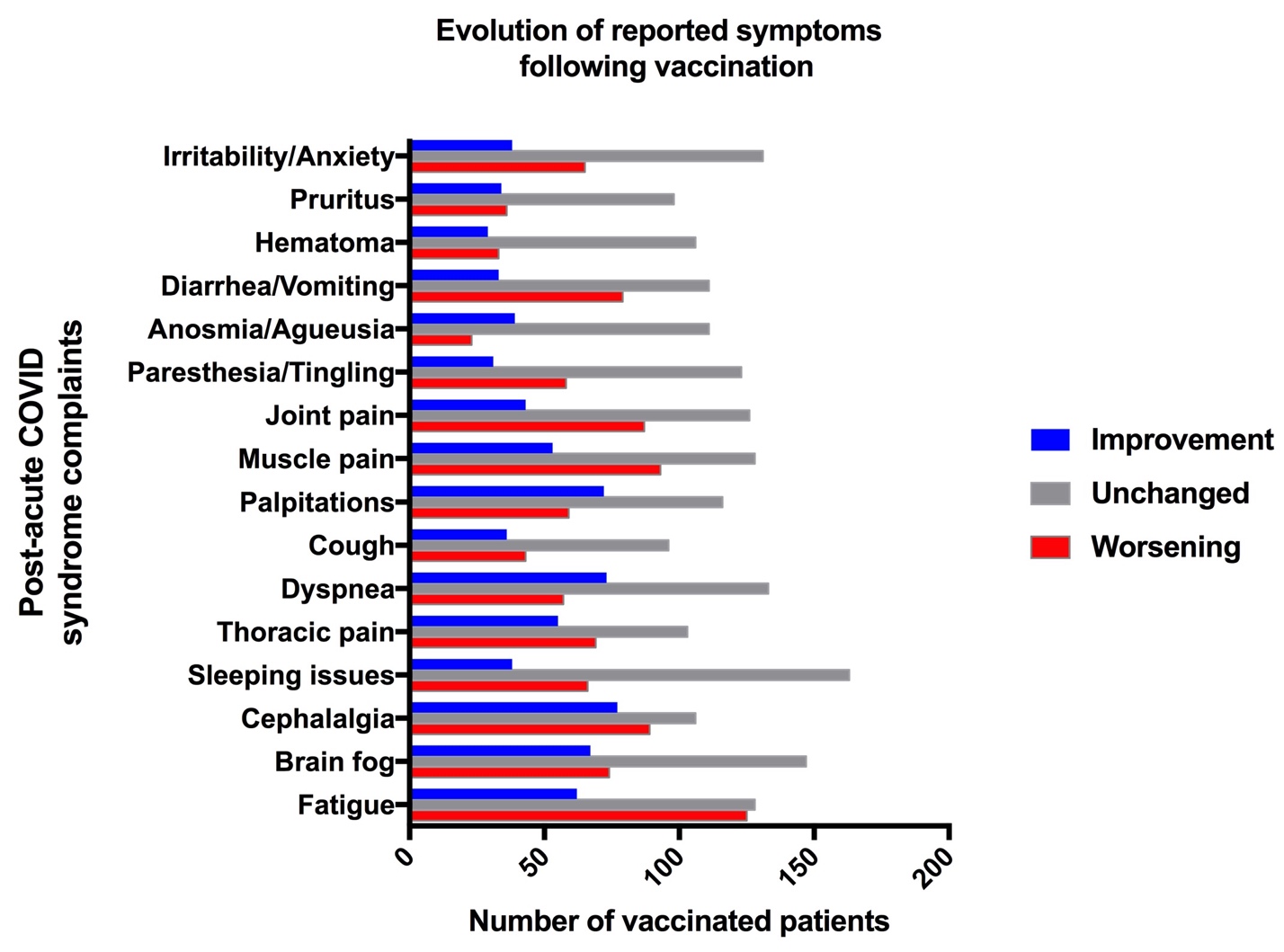

**Supplementary Figure 1. Evolution of reported symptoms following vaccination.**
